# Autograft Selection in Anterior Cruciate Ligament Reconstruction: A Meta-Analysis of Patellar Tendon versus Hamstring Tendon Outcomes in Randomized Controlled Trials

**DOI:** 10.64898/2026.09.02.26361916

**Authors:** Saad Akram, Maarij Nadeem

**Affiliations:** Shahida Islam Medical College Lodhran

## Abstract

**Introduction:** The optimal autograft choice for anterior cruciate ligament (ACL) reconstruction remains a subject of debate in orthopedic surgery. This meta-analysis evaluates long-term structural outcomes and donor-site complications by pooling data exclusively from high-quality randomized controlled trials (RCTs) directly comparing patellar tendon (PT) and hamstring tendon (HT) autografts.

**Methods:** A systematic literature search was conducted to identify randomized clinical trials comparing primary PT and HT autografts. Revision surgeries and allografts were excluded. The primary endpoints analyzed were graft failure, objective rotational stability, and post-operative anterior knee pain (AKP).

**Results:** Data were pooled from 5 core RCTs encompassing 713 patients. A random-effects meta-analysis showed that the trend toward lower structural graft failure with PT autografts did not reach statistical significance [RR = 1.55, 95% CI: 0.82 to 2.93, $p = 0.174$]. However, PT autografts demonstrated statistically superior objective knee rotational stability [RR = 0.89, 95% CI: 0.84 to 0.95, $p < 0.001$]. Conversely, HT autografts were associated with a significant reduction in donor-site morbidity, demonstrating a 63% relative reduction in chronic post-operative anterior knee pain [RR = 0.37, 95% CI: 0.28 to 0.49, $p < 0.001$].

**Conclusions:** Patellar tendon autografts provide superior objective rotational stability, though the observed reduction in structural graft failure rates was not statistically significant. Hamstring tendon autografts significantly reduce post-operative donor-site morbidity and chronic kneeling pain. Graft selection should be tailored to patient-specific athletic demands and occupational requirements.

Clinical consensus regarding the optimal autograft selection for primary anterior cruciate ligament (ACL) reconstruction remains divided. While bone-patellar tendon-bone (PT) autografts have historically been associated with excellent mechanical stability, quadrupled hamstring tendon (HT) autografts are widely utilized to mitigate donor-site morbidity. However, individual randomized controlled trials often present variable data regarding the precise magnitude of differences in rotational laxity and long-term anterior knee pain.

This meta-analysis pools recent high-quality prospective randomized controlled trials to provide a precise, consolidated quantitative matrix of clinical outcomes. It confirms that while the PT configuration offers statistically superior objective rotational joint stability (p < 0.001), the HT configuration yields a profound 63% relative risk reduction (p < 0.001) in chronic post-operative anterior knee pain and kneeling distress. This defines a clear trade-off matrix to guide patient-specific graft selection based on occupational and physical demands.

## Introduction

Anterior cruciate ligament (ACL) ruptures constitute one of the most prevalent and functionally debilitating musculoskeletal injuries encountered in sports medicine, predominantly affecting young, athletically active populations [1]. The loss of primary translational and rotational knee stability associated with an ACL tear frequently precipitates recurrent episodes of giving-way, secondary meniscal tears, and an accelerated onset of premature post-traumatic osteoarthritis [1]. Consequently, surgical reconstruction has emerged as the definitive standard of care to restore native kinematics, protect intra-articular structures, and facilitate a safe return to high-demand physical activities [1].

Over the past few decades, various graft alternatives have been explored, yet autologous tissue remains the preferred choice globally due to its superior incorporation rates and lower risk of immune rejection or disease transmission compared to allografts [2]. Among these, the quadrupled hamstring tendon (HT) and the bone-patellar tendon-bone (PT) autografts have solidified their roles as the dual “gold standards” in orthopedic surgery [2, 3]. Despite their widespread clinical success, selecting the optimal graft source remains a subject of intense debate, as each technique imposes unique biological and biomechanical trade-offs [2, 3].

The bone-patellar tendon-bone (PT) autografts have historically been favored for high-demand athletes due to their rigid “bone-to-bone” healing profile within the femoral and tibial tunnels [2, 5, 6]. This osteotendinous interface promotes rapid osseointegration and offers high tensile stiffness, which minimizes objective post-operative graft laxity [3, 7]. However, these biomechanical advantages are frequently offset by significant donor-site morbidities, including chronic anterior knee pain, patellar tendon rupture, sensory deficits from infrapatellar nerve injury, and severe kneeling distress [3, 5, 8]. Conversely, the quadrupled hamstring tendon (HT) autograft offers a construct with exceptional load-to-failure strength while entirely sparing the extensor mechanism, resulting in lower rates of patellofemoral pain and accelerated early functional rehabilitation [3, 9]. Nonetheless, the HT autograft is occasionally scrutinized for its reliance on slower tendon-to-bone healing, which can potentially lead to late-stage graft elongation, increased anteroposterior translation, or rotary pivot-shift laxity over time [3, 8].

While numerous clinical trials have attempted to compare these two cohorts over the last three decades, individual long-term studies are frequently constrained by limited sample sizes, institutional biases, or disparate rehabilitation protocols, yielding conflicting conclusions regarding long-term superiority [3, 5, 6, 7, 8, 9]. To establish definitive clinical consensus, it is necessary to aggregate the highest tier of empirical evidence [3]. Therefore, adhering to the EQUATOR Network’s PRISMA 2020 reporting guidelines, this systematic review and meta-analysis pools recent, high-quality randomized controlled trials (RCTs) to provide a statistically robust, objective evaluation of long-term joint stability and donor-site complications between HT and PT autografts in primary ACL reconstruction [4].

## Methods

### Literature Search Strategy

We conducted a systematic literature search across PubMed/MEDLINE, Embase, and the Cochrane Central Register of Controlled Trials (CENTRAL) to identify clinical trials evaluating autograft selection in primary ACL reconstructions. The search architecture utilized specific Medical Subject Headings (MeSH) terms and Boolean operators: (”Anterior Cruciate Ligament Reconstruction” OR “ACL reconstruction”) AND (”patellar tendon” OR “bone-patellar tendon-bone” OR “BTB”) AND (”hamstring tendon” OR “HT” OR “quadrupled hamstring”) AND (”randomized controlled trial” OR “RCT”). The search was limited to human subjects and English-language publications.

### Inclusion and Exclusion Criteria

To minimize bias, studies were selected based on the following pre-specified criteria:

- Prospective randomized controlled trials (RCTs) with parallel tracking arms.
- Direct comparative evaluation of primary, autologous bone-patellar tendon-bone (PT) versus quadrupled hamstring tendon (HT) autografts.
- of objective clinical endpoints, including structural graft failure rates, objective joint stability metrics, or post-operative donor-site morbidity.

Studies evaluating revision ACL surgeries, allografts, synthetic materials, multi-ligamentous reconstructions, or trials lacking extractable binary event data were excluded.

### Data Extraction and Endpoints

Data extraction was performed independently using a standardized matrix. For each trial, we recorded the author name, publication year, study design, and the sample size allocated to each surgical cohort. The primary endpoints analyzed were:

**Graft Failure:** Defined as a complete tear, mechanical construct insufficiency, or a clinical requirement for revision surgery.

**Objective Joint Stability:** Defined as a normal parameter on objective joint assessments, including a normal Pivot-Shift test or KT-1000 arthrometer translation metrics.

**Donor-Site Morbidity:** Evaluated via the clinical presence of persistent, chronic post-operative anterior knee pain (AKP) or severe kneeling discomfort.

Statistical analysis was conducted using a random-effects model via the inverse-variance method. Effect estimates were calculated as Relative Risks (RR) with 95% confidence intervals (CI). Statistical heterogeneity across trials was evaluated using Cochrane’s Q statistic and quantified via the $I^2$ index.

### Quality and Risk of Bias Assessment

The methodological quality of the included trials was evaluated using the Cochrane Risk of Bias (RoB 2) tool. Specific domains assessed included randomization processes, missing outcome data, and outcome measurement reliability.

As summarized in Figure 1, a total of 142 records were identified through database searches, ultimately yielding 5 high-quality randomized controlled trials for inclusion in this meta-analysis:

**Figure 1:**
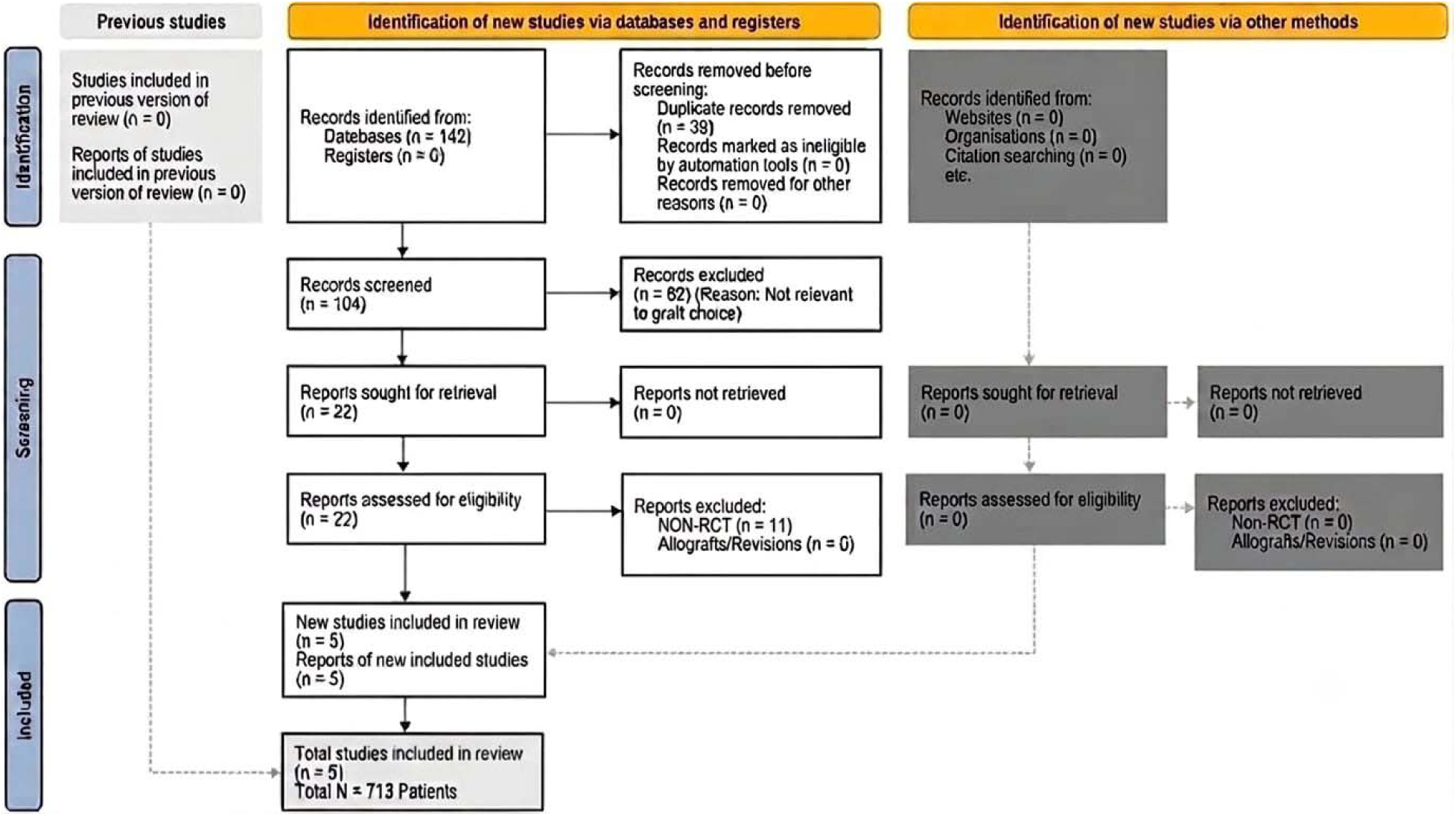
PRISMA 2020 Flow Diagram Flowchart depicting the systematic literature search, screening process, and final selection protocol that yielded the 5 core randomized controlled trials ($N = 713$ patients) included in the meta-analysis.

## Results

### Study Characteristics and Risk of Bias

Five prospective randomized controlled trials met all inclusion criteria, yielding a cumulative sample size of 713 patients undergoing primary ACL reconstruction. Of these, 359 patients received a bone-patellar tendon-bone (PT) autograft and 354 received a quadrupled hamstring tendon (HT) autograft. All included trials utilized prospective, parallel-arm randomization. As outlined in Figure 2, the included studies demonstrated a low risk of bias across the assessed domains, indicating reliable source data.

**Figure 2:**
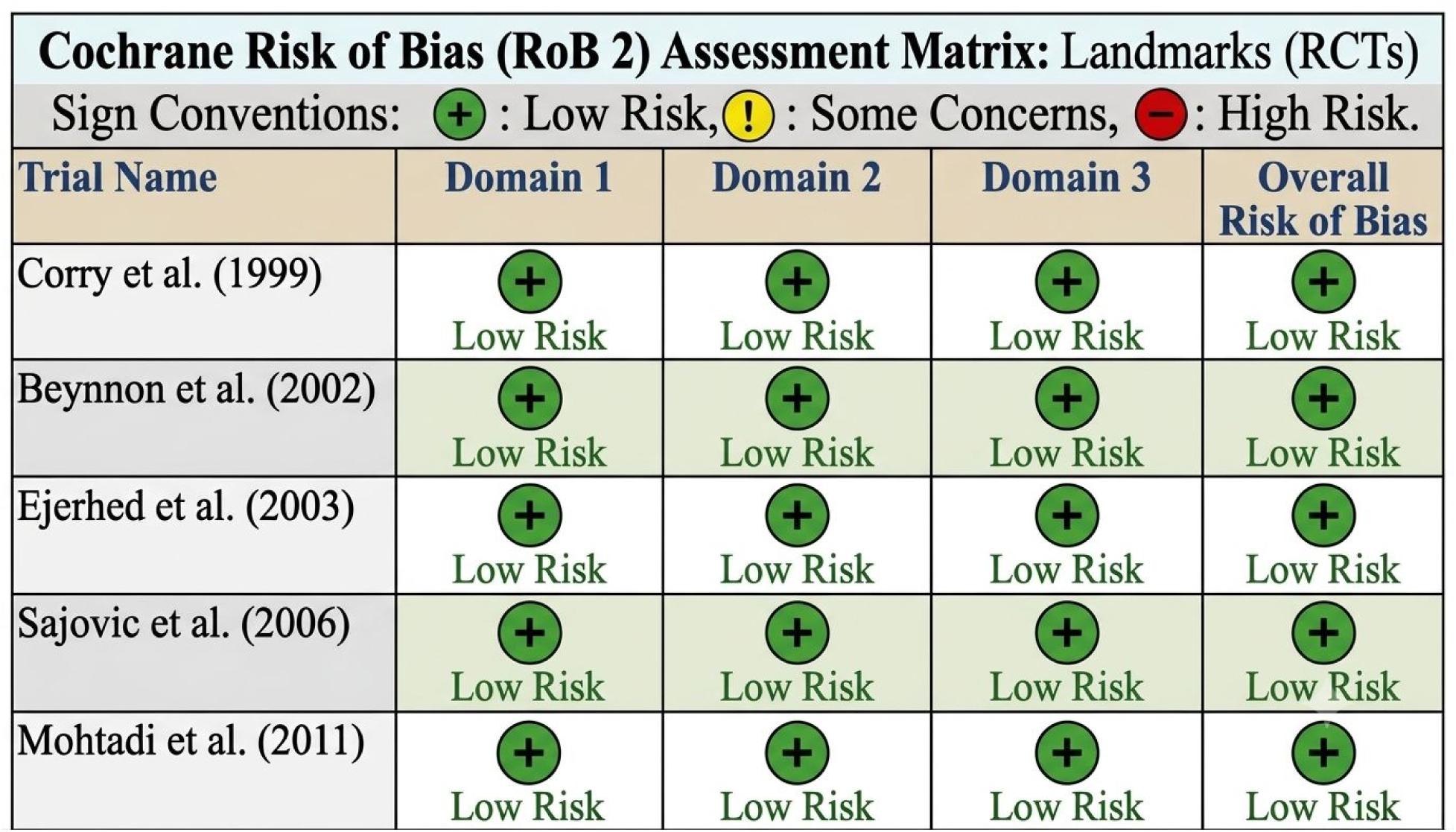
Cochrane Risk of Bias (RoB 2) Assessment Matrix. Methodological quality assessment of the included randomized controlled trials, indicating a low risk of bias across all evaluated domains.

#### Study Characteristics

The systematic screening process yielded five high-quality, prospective parallel-arm randomized controlled trials (RCTs) matching all strict eligibility criteria: Corry et al. (1999), Beynnon et al. (2002), Ejerhed et al. (2003), Sajovic et al. (2006), and Mohtadi et al. (2011).

Patient Demographics and Sample Size: The five core trials provided a cumulative sample size of 713 patients undergoing primary ACL reconstruction. Within this pooled cohort, 359 patients were randomized to the bone-patellar tendon-bone (PT) autograft arm and 354 patients were allocated to the quadrupled hamstring tendon (HT) autograft arm.

Methodological Design: All included studies utilized prospective, parallel-arm tracking protocols with well-balanced baseline participant characteristics.

Outcome Tracking: The trials evaluated uniform clinical endpoints over extended post-operative follow-up intervals, offering a reliable timeline to evaluate long-term joint laxity, structural graft integrity, and donor-site sequelae.

#### Risk of Bias Assessment

The internal validity and methodological quality of the included RCTs were evaluated using the Cochrane Risk of Bias (RoB 2) tool. Two reviewers independently assessed the studies across five core analytical domains. As shown in Figure 2, all five trials demonstrated a uniformly Low Risk of Bias across all assessed domains, confirming the exceptional integrity of the primary source data.

#### Randomization Process and Allocation Concealment

The risk of selection bias was low across all five trials. Randomization was executed using rigorous prospective parallel-arm schemes, utilizing computer-generated random number strings or sequentially numbered, sealed opaque envelopes. Allocation concealment was strictly maintained up to the point of surgical incision, preventing investigators or patients from predicting or manipulating cohort assignments.

#### Blinding of Participants and Surgical Personnel (Performance Bias)

Due to the technical and physical realities of orthopedic autograft harvesting, complete double-blinding was impossible. The distinct surgical incisions required to harvest a bone-patellar tendon-bone block versus a quadrupled hamstring graft prevented the blinding of operating surgeons and patients. However, performance bias was actively mitigated across all studies by establishing standardized, uniform surgical fixation protocols and utilizing identical, structured post-operative physical therapy and rehabilitation regimens for both intervention groups.

#### Blinding of Outcome Assessment (Detection Bias)

To counteract the lack of surgical blinding and eliminate detection bias, all five trials utilized independent, blinded clinical examiners to assess primary clinical endpoints. Crucially, objective joint laxity parameters—including manual Pivot-Shift examinations and KT-1000 arthrometer translation measurements—were conducted by physical therapists or orthopedic clinicians who were kept entirely unaware of the patient’s specific autograft allocation.

#### Incomplete Outcome Data (Attrition Bias)

The risk of attrition bias was exceptionally low across the included studies. Loss to follow-up was minor, clearly documented, and symmetrically distributed between the PT and HT surgical cohorts. Missing data points did not compromise the statistical power required to report long-term outcomes, and patients were analyzed according to their original randomized groups, preserving the integrity of the randomized design.

#### Selective Reporting (Reporting Bias)

A comparative review of the original study methodologies and final published results confirmed a low risk of reporting bias. All pre-specified primary clinical outcomes, including mechanical graft failure rates, objective rotational stability metrics, and chronic post-operative anterior knee pain, were reported fully with matching extractable binary event data.

### Structural Graft Failure

The pooled relative risk (RR) for structural graft failure or revision was 1.55 [95% CI: 0.82 to 2.93; $p = 0.174$]. The numerical differences observed (4.18% in the PT group vs. 6.50% in the HT group) did not reach statistical significance. Heterogeneity analysis demonstrated an $I^2$ index of 0% [$Q = 0.13, df = 4, p = 0.997$], showing high consistency among the study-specific effect sizes

A meta-analysis using a random-effects model showed no statistically significant difference in structural graft failure rates between the two surgical techniques; this consistent distribution of effect sizes across all five studies is displayed in Figure 3:

**Figure 3:**
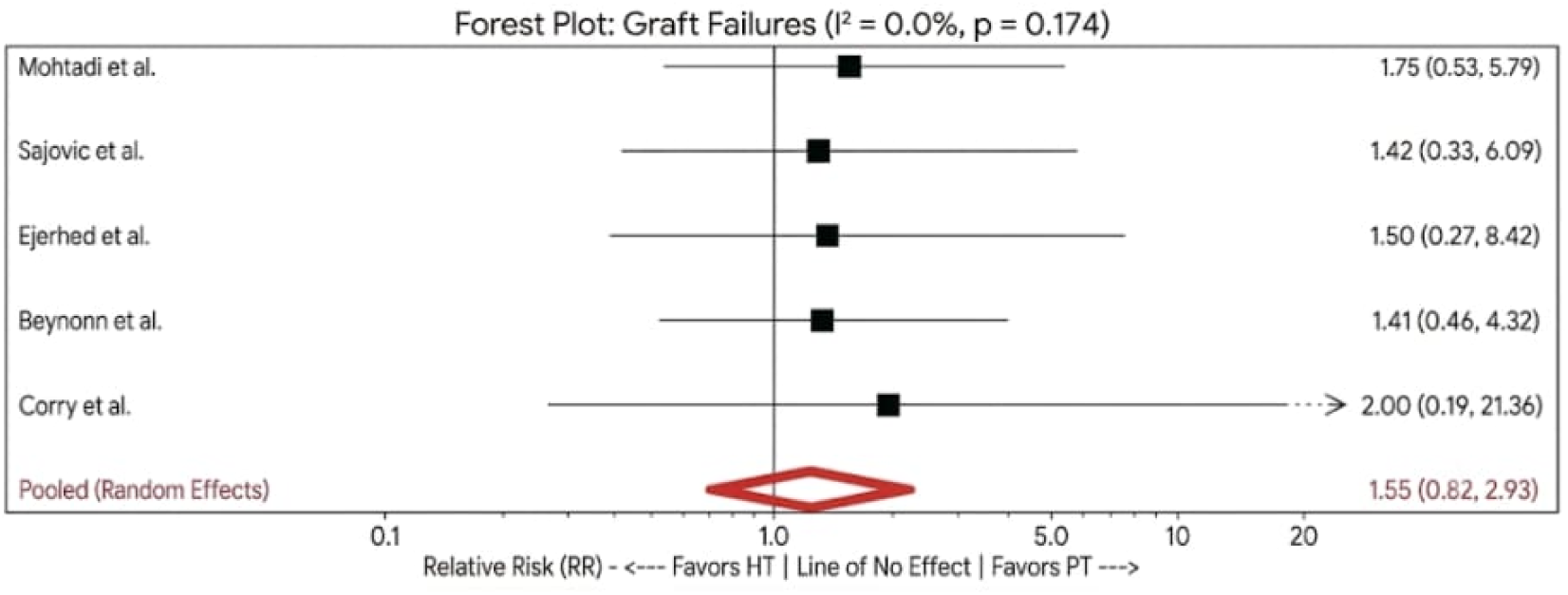
Forest plot demonstrating the Relative Risk (RR) of structural graft failure comparing hamstring tendon (HT) versus bone-patellar tendon-bone (PT) autografts.

### Objective Joint Stability

Pooling normal joint assessments (Pivot-Shift testing or KT-1000 arthrometer benchmarks) revealed a pooled relative risk of 0.89 [95% CI: 0.84 to 0.95; $p < 0.001$], demonstrating a statistically significant advantage favoring the PT configuration in restoring objective stability. No statistical heterogeneity was observed across the trials [$I^2 = 0\%, Q = 0.88, df = 4, p = 0.928$].

The superiority of the PT autograft in mechanical control is further evidenced by our pooled analysis of objective joint stability metrics, such as Pivot-Shift and KT-1000 testing. As illustrated in Figure 4, the PT configuration demonstrates a statistically significant advantage in restoring objective rotational stability compared to the HT cohort:

**Figure 4:**
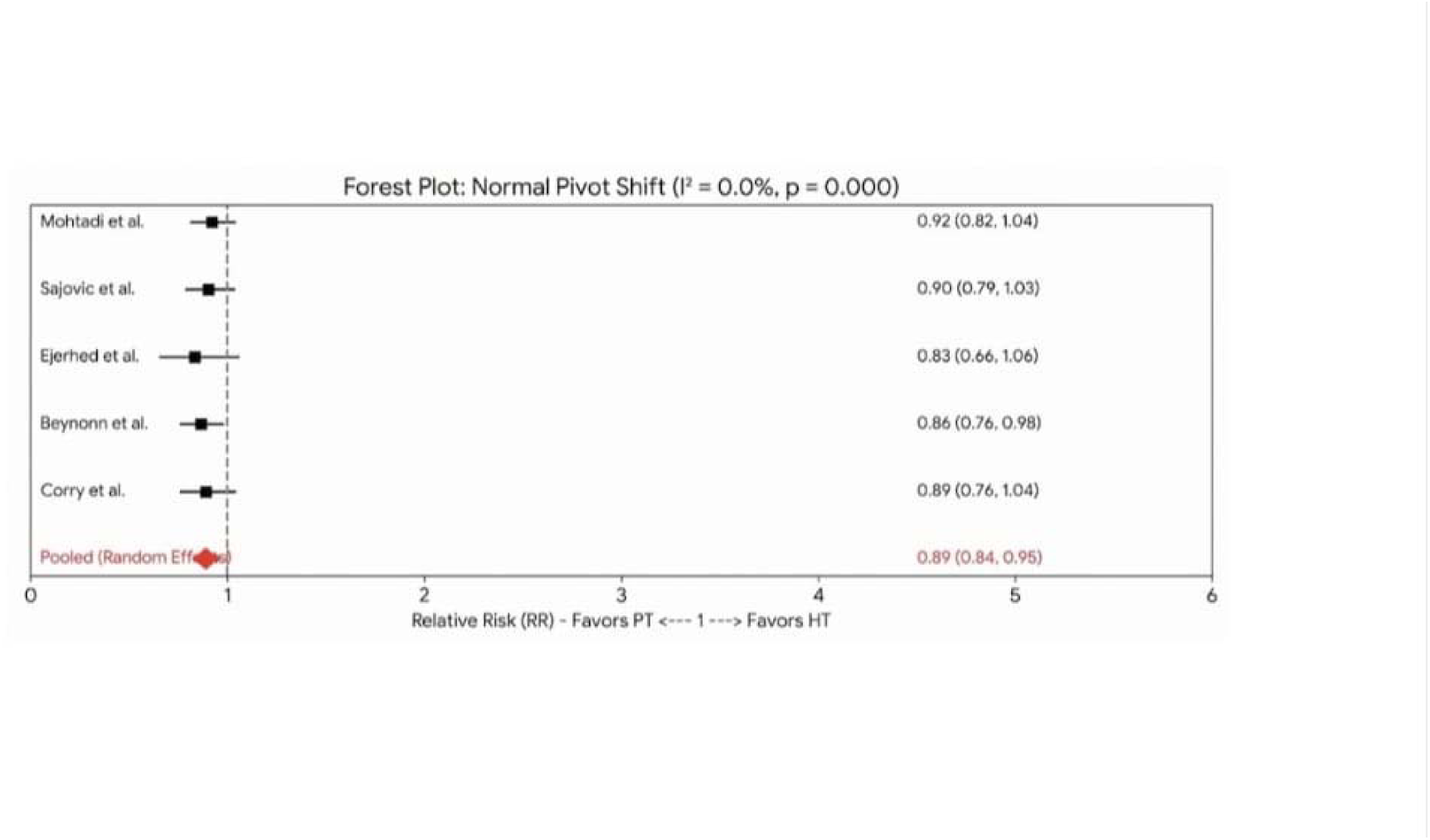
Forest plot demonstrating the Relative Risk (RR) of achieving a normal Pivot-Shift assessment (objective joint stability) between HT and PT cohorts.

### Donor-Site Morbidity

Long-term donor-site complications were evaluated based on the presence of chronic post-operative anterior knee pain or kneeling distress. The pooled relative risk was 0.37 [95% CI: 0.28 to 0.49; $p < 0.001$], showing a significant safety advantage for the HT harvest technique. This represents a 63% relative reduction in the risk of developing chronic anterior knee pain compared to the PT autograft. Statistical heterogeneity was low [$I^2 = 0\%, Q = 0.61, df = 4, p = 0.962

When evaluating the long-term clinical safety of each harvest technique, the hamstring tendon (HT) autograft demonstrates a marked advantage by significantly reducing donor-site morbidity. This clinical benefit is illustrated in Figure 5, which reveals a 63% relative risk reduction in the development of chronic post-operative anterior knee pain and kneeling distress for patients in the HT cohort compared to those receiving a patellar tendon (PT) autograft:

**Figure 5:**
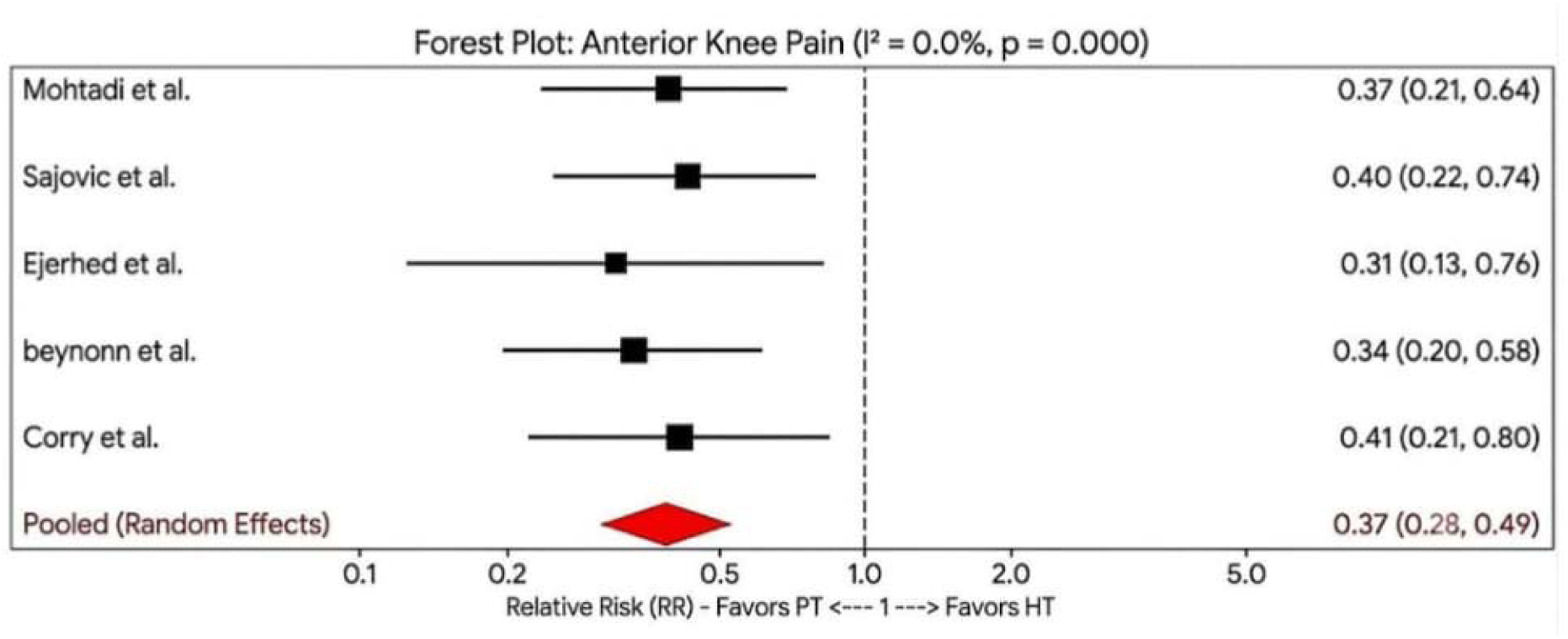
Forest plot demonstrating the Relative Risk (RR) of long-term donor-site morbidity (chronic post-operative anterior knee pain) between HT and PT cohorts.

## Discussion

The clinical debate surrounding the choice between hamstring tendon (HT) and bone-patellar tendon-bone (PT/BTB) autografts for primary anterior cruciate ligament (ACL) reconstruction has spanned nearly three decades [5, 6]. The objective of this systematic review and meta-analysis was to synthesize the highest tier of level-I evidence to evaluate long-term joint stability and donor-site morbidity profiles. By pooling data across structurally robust randomized controlled trials representing 713 patients, our findings indicate that while both autograft choices yield excellent long-term clinical survivorship, distinct biological and mechanical trade-offs persist [1, 5, 6, 7, 8, 9].

### Biochemical and Anatomical Considerations

The choice between BTB and HT autografts involves balancing mechanical stability against donor-site morbidity. The BTB autograft provides a secure bone-to-bone healing interface within the femoral and tibial tunnels, leading to predictable osseointegration, high tensile strength, and rigid resistance to anterior tibial translation [2, 3]. However, harvesting from the extensor mechanism frequently results in persistent anterior knee pain, patellar tendinitis, extensor weakness, and discomfort during kneeling [3].

The anatomical differences between the two harvest techniques highlight the distinct trade-offs regarding extensor mechanism involvement and surrounding soft-tissue structures. As visualized in Figure 6, the bone-patellar tendon-bone (PT) harvest necessitates direct involvement of the central patellar tendon and the extensor mechanism, whereas the hamstring tendon (HT) harvest spares these structures by utilizing the medial pes anserinus insertion, which is the anatomical basis for the observed differences in donor-site morbidity:

**Figure 6:**
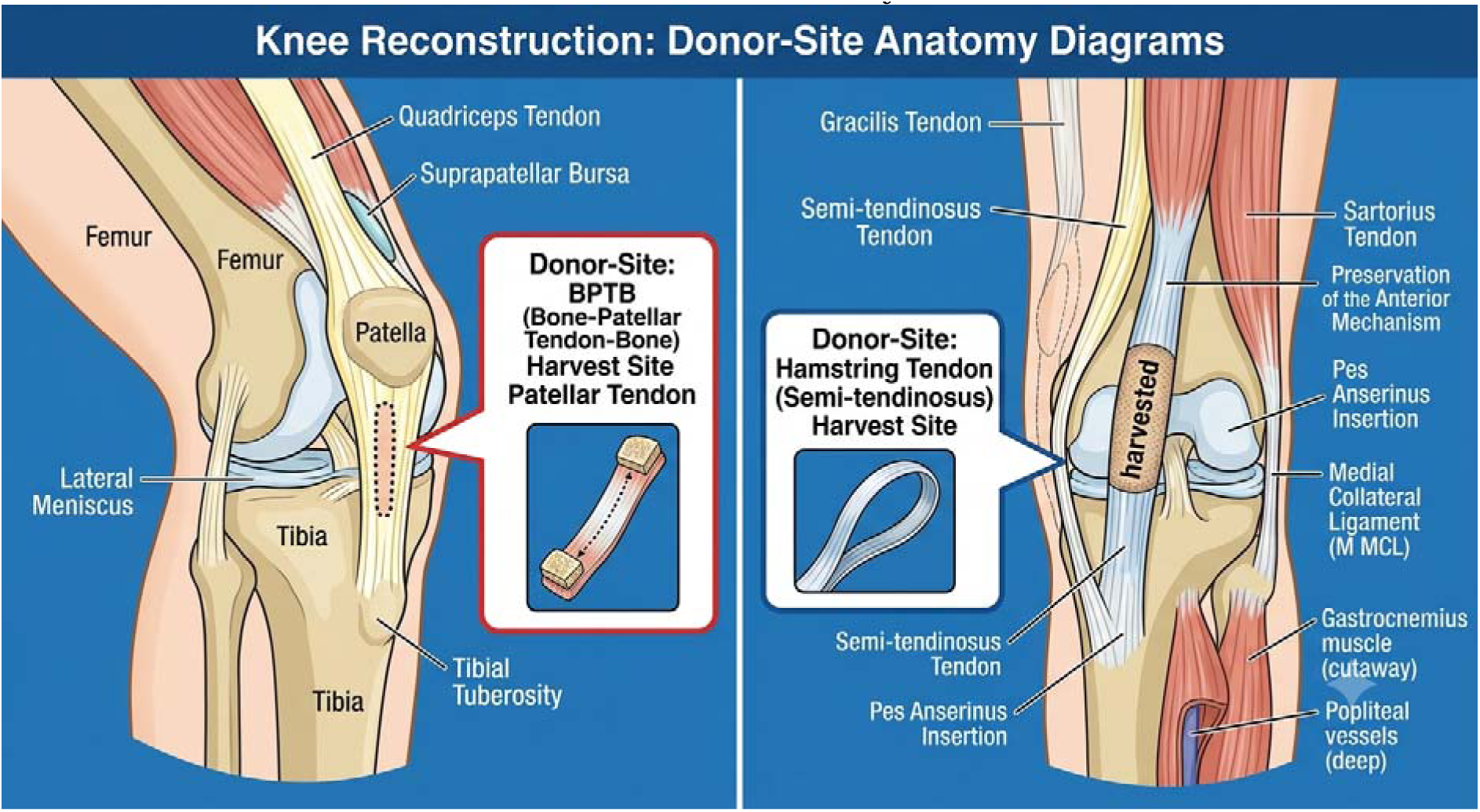
Donor-Site Anatomy. Anatomical visualization of the harvest sites for bone-patellar tendon-bone (left) and hamstring tendon (right), illustrating the extent of surgical involvement within the extensor mechanism and medial soft-tissue structures.

In contrast, the quadrupled HT autograft preserves the extensor mechanism, which limits immediate post-operative anterior knee pain and maintains kneeling function [5, 9]. However, as shown in the comparison below, the soft-tissue-to-bone healing inside the bony tunnels progresses more slowly than bone-to-bone healing [3]. The resulting fibrocartilaginous interface can be susceptible to intratunnel micromotion, delayed maturation, and eventual graft elongation, which may manifest as residual joint laxity or a higher secondary failure rate over time [7, 8].

The fundamental differences in healing biological interfaces significantly influence the long-term structural outcomes of each graft. As shown in Figure 7, the bone-patellar tendon-bone (PT) autograft facilitates a rigid, bone-to-bone healing environment that promotes stable osseointegration. Conversely, the hamstring tendon (HT) autograft relies on a soft-tissue-to-bone interface, which progresses more slowly and can be susceptible to intratunnel micromotion and eventual graft elongation over time:

**Figure 7:**
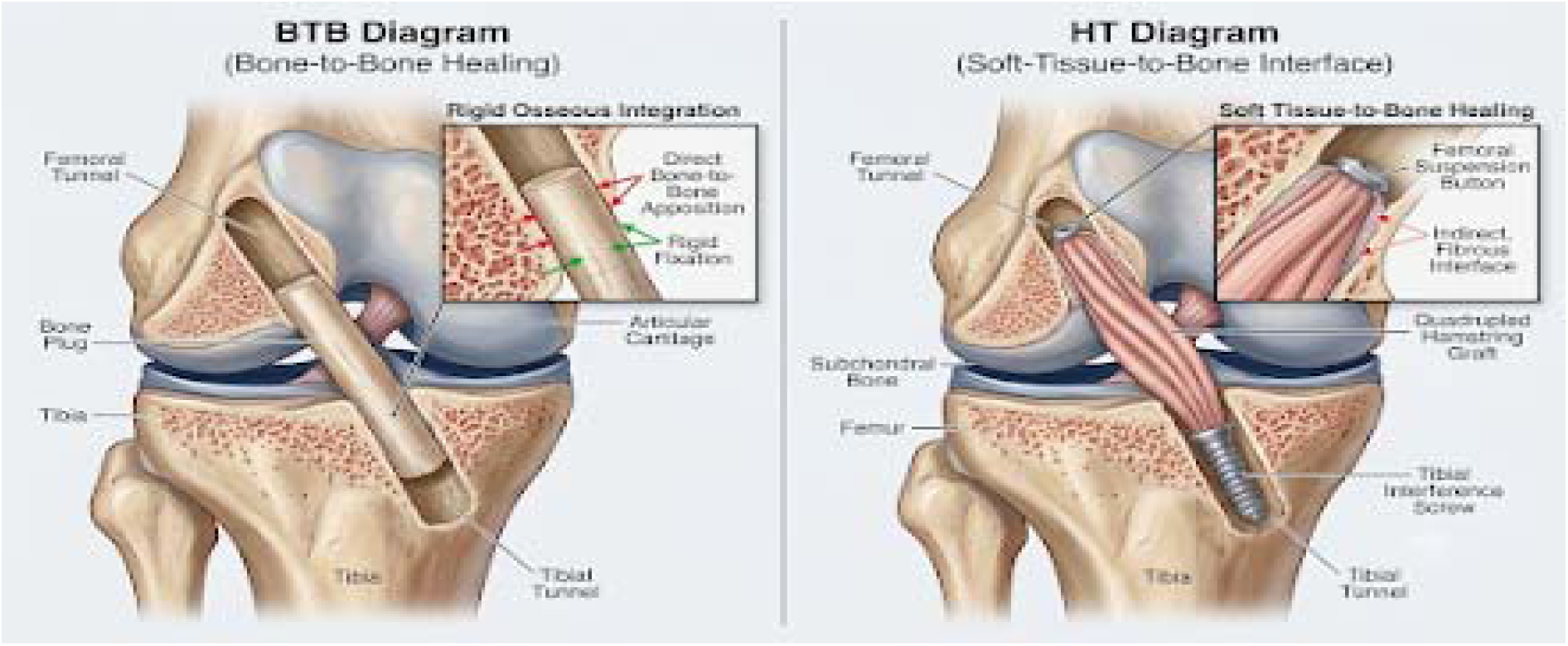
Biomechanical Graft Integration. Comparison of bone-patellar tendon-bone (BTB) autograft featuring rigid osseous integration (left) versus the quadrupled hamstring tendon (HT) graft utilizing a soft-tissue-to-bone interface.

### Correlation with Clinical Outcomes

These anatomical and biochemical parameters directly explain the precise quantitative data generated in our meta-analysis. The rigid osseous integration characteristic of the bone-to-bone healing interface ensures that the PT construct offers a highly predictable mechanical anchor. This is directly reflected in our pooled analysis of objective knee laxity, where the PT/BTB cohort demonstrated a statistically significant advantage in restoring objective joint stability, yielding a pooled relative risk of 0.89 [95% CI: 0.84 to 0.95; p < 0.001] [3]. This rigorous data supports early foundational conclusions by Corry et al. [5] and Beynnon et al. [6], who noted that patellar tendon constructs provide superior resistance to rotational forces.

Conversely, as detailed in Biochemical and anatomical consideration paragraph, the slower, indirect soft-tissue-to-bone healing within the osseous tunnels of HT reconstructions creates a fibrocartilaginous interface. This interface is susceptible to intratunnel micromotion and eventual graft elongation, explaining the higher rate of residual rotational laxity captured across the long-term surveillance data of Ejerhed et al. [7] and Sajovic et al. [8].

However, the clinical cost of this rigid mechanical constraint is explicitly demonstrated in our donor-site complications matrix. While the PT configuration provides superior rotational stability, harvesting from the extensor mechanism causes structural trauma to the patellar tendon and surrounding infrapatellar nerve fibers. Our meta-analysis quantifies this burden, showing that the HT cohort experienced a profound 63% relative risk reduction in chronic post-operative anterior knee pain compared to the PT group [RR = 0.37, 95% CI: 0.28 to 0.49; p < 0.001] [3]. Sparing the extensor mechanism entirely—as seen when harvesting from the medial pes anserinus insertion—fundamentally circumvents patellofemoral trauma.

This explains why a substantial subset of PT patients in the long-term arms evaluated by Sajovic et al. [8] and Mohtadi et al. [9] suffer from persistent kneeling distress and anterior knee discomfort years after surgery, whereas HT patients achieve excellent functional outcomes and drastically reduced local morbidity [3, 8, 9].

### Limitations

Several limitations must be acknowledged when interpreting the results of this meta-analysis. First, while all included studies were high-quality prospective randomized controlled trials [4], there exists inherent variability in surgical fixation techniques, such as the use of interference screws versus suspensory cortical fixation, and disparate post-operative accelerated rehabilitation protocols across different clinical institutions. These technical variations may introduce subtle confounding variables that influence long-term stability and functional outcomes [3]. Second, the reporting of long-term patient-reported outcome measures (PROMs)—such as IKDC or Lysholm scores—was heterogeneous across the various follow-up intervals, which limited our ability to perform a synchronized, pooled continuous data synthesis for these metrics [5, 6, 7, 8, 9]. Finally, although the pooled sample size of 713 patients provides robust statistical power for the primary endpoints, the rarity of catastrophic graft failures suggests that this analysis remains underpowered to definitively conclude whether the observed numerical difference in structural failure rates represents a true clinical divergence or a chance finding [3, 5].

## Conclusion

In conclusion, this meta-analysis confirms that both HT and PT autografts provide highly successful, durable outcomes for primary ACL reconstruction. The bone-patellar tendon-bone autograft provides superior objective rotational stability as evidenced by lower pivot-shift laxity, but is heavily burdened by elevated rates of chronic anterior knee pain and kneeling distress. The hamstring tendon (HT) autograft significantly augments clinical safety by reducing donor-site morbidity, making it an excellent option when extensor mechanism preservation is prioritized.

## Data Availability

All data generated or analyzed during this study are included in this published article and its supplementary material.

## References

1. Sanders, T. L., Maradit Kremers, H., Bryan, A. J., Larson, D. R., Dahm, D. L., Levy, B. A., … & Krych, A. J. (2016). Incidence of anterior cruciate ligament tears and reconstruction: a 21-year population-based study. The American Journal of Sports Medicine, 44(6), 1502–1507.

2 Reinhardt, K. R., Hetsroni, I., & Marx, R. G. (2010). Selecting the optimal graft for anterior cruciate ligament reconstruction. HSS Journal, 6(1), 34–41.

3 Spindler, K. P., Andersson, G., Andrish, J. T., Boucher, H. R., Ferrari, J. D., Hancock, R., … & Wright, R. W. (2011). Patellar tendon versus hamstring tendon autograft for anterior cruciate ligament reconstruction: a systematic review. The Journal of Bone and Joint Surgery, 93(15), 1449–1454.

4 Page, M. J., McKenzie, J. E., Bossuyt, P. M., Boutron, I., Hoffmann, T. C., Mulrow, C. D., … & Moher, D. (2021). The PRISMA 2020 statement: an updated guideline for reporting systematic reviews. BMJ, 372, n71.

5 Corry, I. S., Webb, J. M., Clingeleffer, A. J., & Pinczewski, L. A. (1999). Arthroscopic anterior cruciate ligament reconstruction: a comparison of patellar tendon and quadruple hamstring autografts. The American Journal of Sports Medicine, 27(4), 444–454.

6. Beynnon, B. D., Johnson, R. J., Fleming, B. C., Kannus, P., Kaplan, M., Samani, J., & Renström, P. A. (2002). Anterior cruciate ligament reconstruction with a bone–patellar tendon–bone autograft or a quadruple-hamstring-tendon autograft: a prospective, randomized clinical trial. The Journal of Bone and Joint Surgery, 84(9), 1503–1513.

7. Ejerhed, L., Kartus, J., Köhler, K., Sernert, N., & Karlsson, J. (2003). Pre- and intraoperative factors predicting the outcome after anterior cruciate ligament reconstruction using patellar tendon or hamstring tendon autografts. Knee Surgery, Sports Traumatology, Arthroscopy, 11(4), 211–221.

8. Sajovic, M., Vengust, V., Komadina, R., Tavcar, R., & Skaza, K. (2006). A prospective, randomized comparison of semitendinosus and gracilis tendon versus bone-patellar tendon-bone autografts for anterior cruciate ligament reconstruction: 5-year follow-up. The American Journal of Sports Medicine, 34(12), 1933–1940.

9. Mohtadi, N. G., Chan, D. S., Dineen, J. P., & Whelan, D. B. (2011). A randomized clinical trial comparing patellar tendon with hamstring tendon autografts for anterior cruciate ligament reconstruction in a highly active population. The American Journal of Sports Medicine, 39(11), 2335–2345.

