## supplemental file 2 for "Autograft Selection in Anterior Cruciate Ligament Reconstruction: A Meta-Analysis of Patellar Tendon versus Hamstring Tendon Outcomes in Randomized Controlled Trials"

**PRISMA-checklist**

| **Subjects** | **PRISMA for ACL Reconstruction** |
| --- | --- |
| ***Title*** |  |
| **Title** | Autograft Selection in Anterior Cruciate Ligament Reconstruction: A Meta-Analysis of Patellar Tendon versus Hamstring Tendon Outcomes in Randomized Controlled Trials |
| ***Abstract*** |  |
| **Structured summary** | Provided in the text with Background, Methods, Results, and Conclusions sections. (Total N = 713 patients across 5 core RCTs; No registration number reported). |
| ***Introduction*** |  |
| **Rationale** | Clinical consensus on optimal autograft choice is divided. Bone-patellar tendon-bone (PT) provides mechanical stability but causes donor-site morbidity, while quadrupled hamstring tendon (HT) mitigates donor-site pain but has variable data regarding rotational laxity. |
| **Objectives** | Direct comparison of long-term structural outcomes, joint stability, and donor-site morbidity of bone-patellar tendon-bone versus quadrupled hamstring tendon autografts utilizing strictly prospective RCTs |
| ***Methods*** |  |
| **Protocol and registration** | This systematic review and meta-analysis was not registered in a database such as PROSPERO. No formal protocol was published prior to the initiation of this study. |
| **Eligibillity Criteria** | Inclusion criteria: Peer-reviewed randomized controlled trials (RCTs) comparing primary bone-patellar tendon-bone (PT) autografts versus quadrupled hamstring tendon (HT) autografts for anterior cruciate ligament reconstruction (ACLR). Studies were required to report at least one primary clinical outcome (e.g., structural graft failure, objective rotational stability via Pivot-Shift, or donor-site anterior knee pain) with a minimum follow-up of 24 months.  Exclusion criteria: Non-randomized cohorts, retrospective studies, revision ACLRs, multiligament injuries, or use of allografts/synthetic grafts. |
| **Information Sources** | PubMed/MEDLINE, Embase, and the Cochrane Central Register of Controlled Trials (CENTRAL). Last search date and coverage dates not explicitly reported. |
| **Search** | ("Anterior Cruciate Ligament Reconstruction" OR "ACL reconstruction") AND ("patellar tendon" OR "bone-patellar tendon-bone" OR "BTB") AND ("hamstring tendon" OR "HT" OR "quadrupled hamstring") AND ("randomized controlled trial" OR "RCT"). |
| **Study selection** | Multi-stage process: 142 records identified ; 38 duplicates removed ; 104 screened by title/abstract ; 82 excluded ; 22 full-text articles assessed ; 17 excluded (11 non-RCT, 6 allografts/revisions) ; 5 core RCTs included. |
| **Data collection**  **Process** | Data extraction performed independently using a standardized matrix. |
| **Data items** | Extracted variables included study characteristics (author, publication year, country, sample size, follow-up duration), participant demographics (age, sex), graft specifications (bone-patellar tendon-bone vs. quadrupled hamstring tendon), and primary clinical outcomes (graft failure rates, Pivot-Shift objective stability, and post-operative anterior knee pain). |
| **Risk of bias in**  **individual studies** | The risk of bias within individual studies was evaluated using the Cochrane Risk of Bias (RoB 2) tool. Two independent reviewers assessed each included randomized controlled trial across the following domains: randomization process, deviations from intended interventions, missing outcome data, measurement of the outcome, and selection of the reported result. Each study was categorized as having 'Low Risk,' 'Some Concerns,' or 'High Risk' of bias. Discrepancies between reviewers were resolved through consensus. As shown in Figure 2, all 5 core RCTs were determined to be at 'Low Risk' across all assessed domains. |
| **Summary measures** | Relative Risks (RR) with 95% confidence intervals (CI). |
| **Synthesis of results** | Random-effects model via the inverse-variance method. Statistical heterogeneity evaluated using Cochrane's Q statistic and quantified via the I^2 index. |
| **Risk of bias across**  **studies** | The potential for risk of bias across studies, specifically publication bias, was investigated. Given that the number of included randomized controlled trials was fewer than ten, we utilized funnel plot analysis to visually assess for asymmetry. We did not employ formal statistical tests for funnel plot asymmetry (e.g., Egger’s or Begg’s test) because such methods have limited power to distinguish between real bias and chance when the number of studies is small. |
| **Additional analyses** | No pre-specified subgroup or sensitivity analyses were performed for this meta-analysis due to the limited number of eligible randomized controlled trials. |
| ***Results*** |  |
| **Study selection** | Described in text and flowchart format: 142 records identified ; 104 screened ; 22 full-text assessed ;5 core RCTs included (N = 713 total patients: 359 PT, 354 HT) |
| **Study characteristics** | 5 included trials spanning 1999–2011: Corry (1999), Beynnon (2002), Ejerhed (2003), Sajovic (2006), Mohtadi (2011). |
| **Risk of bias within**  **studies** | Figure2 indicates "Low Risk" across all assessed domains (Randomization, Missing Data, Outcome Measurement, and Overall Risk) for all 5 studies. |
| Results of individual  studies | Individual RRs and CIs are provided inside the Forest Plots (Figures 1, 2, and 3) for each of the 5 trials. |
| **Synthesis of results** | Structural Graft Failure: [RR = 1.55, 95% CI: 0.82 to 2.93, p = 0.174; I^2 = 0%] (Not statistically significant). |
| **Risk of bias across**  **studies** | Risk of bias across studies was assessed visually using funnel plots. Due to the limited number of included studies (n=5), formal statistical testing for funnel plot asymmetry (e.g., Egger’s test) was not conducted, as it lacks sufficient power to distinguish chance from real bias. |
| **Additional analysis** | No sensitivity or subgroup analyses were performed in this review. The small number of included studies and the consistency in primary outcomes across the randomized controlled trials (RCTs) precluded further sub-stratification. |
| ***Discussion*** |  |
| **Summary of evidence** | Confirming trade-off matrix: PT autografts yield statistically superior objective rotational joint stability (p < 0.001). However, HT configurations offer a profound 63% relative risk reduction (p < 0.001) in chronic post-operative anterior knee pain and kneeling distress. |
| **Limitations** | Included trials span multiple decades (varied surgical fixation technologies); variations in post-operative rehab protocols; subjectively quantified pain thresholds for anterior knee pain. |
| **Conclusions** | Autograft selection should not be protocol-driven but tailored individually based on the patient's athletic demands (favoring PT for high-demand pivoting) versus occupational/lifestyle requirements (favoring HT for runners, mechanics, or activities requiring kneeling) |
| **Funding** |  |
| **Funding** | This research received no specific grant from any funding agency in the public, commercial, or not-for-profit sectors. |

Note: Item 1 (Title): Title Page.

Item 3 (Rationale): Introduction (under "Clinical Context" and "Biomechanical and Anatomical Considerations").

Item 4 (Objectives): Introduction (under "Objectives").

Item 6 (Eligibility criteria): Methods (under "Inclusion and Exclusion Criteria").

Item 8 (Search): Methods (under "Literature Search Strategy").

Item 12 (Risk of bias individual): Methods (under "Quality and Risk of Bias Assessment").

Item 17 (Study selection): Results (under "Study Characteristics and Risk of Bias" and Figure 1 flowchart).

Item 21 (Synthesis of results): Results (under "Structural Graft Failure", "Objective Joint Stability", and "Donor-Site Morbidity").
